# Machine learning quantification of amyloid deposits in histological images of ligamentum flavum

**DOI:** 10.1101/2021.12.05.21267317

**Authors:** Andy Y. Wang, Vaishnavi Sharma, Harleen Saini, Joseph N. Tingen, Alexandra Flores, Diang Liu, Mina G. Safain, James Kryzanski, Ellen D. McPhail, Knarik Arkun, Ron I. Riesenburger

**Affiliations:** Department of Neurosurgery, Tufts Medical Center, Boston, Massachusetts, 02111, USA; Department of Laboratory Medicine and Pathology, Mayo Clinic, Rochester, Minnesota, 55905 USA; Department of Pathology, Tufts Medical Center, Boston, Massachusetts, 02111, USA

**Keywords:** Wild-type transthyretin amyloid, Ligamentum flavum, Trainable Weka Segmentation, Machine learning, Color thresholding

## Abstract

**Background:** Wild-type transthyretin amyloidosis (ATTRwt) is an underdiagnosed and potentially fatal disease. Interestingly, ATTRwt deposits have been found to deposit in the ligamentum flavum (LF) of patients with lumbar spinal stenosis prior to the development of systemic and cardiac amyloidosis. In order to study this phenomenon and its possible relationship with LF thickening and systemic amyloidosis, a precise method of quantifying amyloid deposits in histological slides of LF is critical. However, such a method is currently unavailable. Here, we present a machine learning quantification method with Trainable Weka Segmentation (TWS) to assess amyloid deposition in histological slides of LF.

**Methods:** Images of ligamentum flavum specimens stained with Congo red are obtained from spinal stenosis patients undergoing laminectomies and confirmed to be positive for ATTRwt. Amyloid deposits in these specimens are classified and quantified by TWS through training the algorithm via user-directed annotations on images of LF. TWS can also be automated through exposure to a set of training images with user-directed annotations, and then application to a set of new images without additional annotations. Additional methods of color thresholding and manual segmentation are also used on these images for comparison to TWS.

**Results:** We develop the use of TWS in images of LF and demonstrate its potential for automated quantification. TWS is strongly correlated with manual segmentation in the training set of images with user-directed annotations (R = 0.98; p = 0.0033) as well as in the application set of images where TWS was automated (R = 0.94; p = 0.016). Color thresholding was weakly correlated with manual segmentation in the training set of images (R = 0.78; p = 0.12) and in the application set of images (R = 0.65; p = 0.23).

**Conclusion:** TWS machine learning closely correlates with the gold standard comparator of manual segmentation and outperforms the color thresholding method. This novel machine learning method to quantify amyloid deposition in histological slides of ligamentum flavum is a precise, objective, accessible, high throughput, and powerful tool that will hopefully pave the way towards future research and clinical applications.

## BACKGROUND

Wild-type transthyretin amyloidosis is likely an underdiagnosed disease that has been studied in the context of cardiac amyloidosis and carpal tunnel syndrome [1–3]. Lumbar spinal stenosis has been increasingly indicated as a potential early manifestation of amyloidosis, with ATTRwt found to deposit in the ligamentum flavum (LF) of spinal stenosis patients [3–5]. Previously, our group has shown that spinal stenosis patients harboring wild-type transthyretin amyloid in histological samples of LF have thicker LF than patients without amyloid [6,7]. To enable further research into understanding the possible relationship of amyloid deposition with LF thickening and systemic amyloidosis, a method to quantify the precise amount of amyloid in histological specimens of LF is necessary. However, no optimal technique currently exists. The focus of this paper is to present a novel, objective, and accurate machine learning method of quantifying amyloid deposition in histological LF specimens.

Previous studies have attempted to quantify amyloid deposition in ligamentous tissue, but methods thus far have not been adequately precise, reproducible, and feasible. Some studies used qualitative methods based on visual inspection of specimens and grouping them into broad categories of amyloid load [8,9]. One such study involving tenosynovial tissue from carpal tunnel release surgery qualitatively grouped amyloid deposition into three grades of “mild”, “moderate”, and “severe” [10]. More advanced quantification methods utilize computer software such as ImageJ to analyze digitally scanned histology images. One study used ImageJ to quantify amyloid deposits in the ligamentum flavum, but did not specify details [5]. Another study used the color thresholding function in ImageJ with detailed descriptions, which we attempted to replicate [11]. However, we encountered notable shortcomings of this method, such as significant overestimation or underestimation of amyloid load, and the need to manually adjust thresholding values for each scan due to inherent color variations from Congo red staining. Ideally, the gold standard is for an experienced neuropathologist to manually circumscribe regions of amyloid deposits on digital scans, but this process is too time-consuming for practical use. While the above studies have attempted to quantify amyloid deposition in LF, their methods remain qualitative, not clearly described, or imprecise.

We explore the use of Trainable Weka Segmentation, a machine learning algorithm plugin within Fiji/ImageJ, to quantify and analyze amyloid deposits in histological slides of ligamentum flavum [12,13]. Notably, the TWS method enables analysis of microscope and histology images through user-directed annotations that train the algorithm to recognize certain components of an image [12]. TWS learns from a limited number of manual annotations and classifies the rest of the pixels of the image into different classes of interest. The use of this algorithm arose from basic science research where it has been used in studies to count colonies and track cells, analyze stained mouse muscle sections, and recognize amyloid plaques in a mouse model of Alzheimer’s disease [14–16]. In clinical research, this method has been emerging in various uses, such as to analyze human CT scans and to delineate boundaries of breast carcinoma in human tissue [17,18]. Here, we describe the first use of the TWS machine learning algorithm to quantify amyloid deposits in histological samples of ligamentum flavum from patients with spinal stenosis.

## METHODS

### Histological images of ligamentum flavum

An IRB-approved cohort of spinal stenosis patients undergoing laminectomies had ligamentum flavum samples dissected and sent to pathology. Formalin-fixed, paraffin-embedded ligamentum flavum sections were stained with Congo red and visualized under polarized light to identify amyloid deposits, then typed by mass spectrometry (Mayo Clinic Laboratories, Rochester, MN, USA). All specimens used in this study were positive for ATTRwt per mass spectrometry-based proteomics. Ten slides of ligamentum flavum sections were randomly selected, scanned, and digitized using the VENTANA DP 200 slide scanner (Roche Diagnostics, Rotkreuz, Switzerland) at a 20x magnification and 1 focus layer.

### Trainable Weka Segmentation

Trainable Weka Segmentation (TWS) is a plugin that can be accessed within Fiji, a variation of ImageJ (Version 2.1.0/1.53c) with more extensive biology-specific plugins including Bio-Formats (Figure 1A). Feature settings are selected to include gaussian blur, difference of gaussians, and Sobel operator. Four different tissue components are chosen as classes: amyloid, glass slide, calcifications/bone dust, and ligamentum flavum tissue (Figure 1A). A LF image is opened within the TWS graphical user interface, and a few manual annotations corresponding to each class of interest is then drawn on the image (Figure 1B). “Train classifier” initiates a process in which TWS attempts to learn from these annotations and segments the rest of the image into the preselected classes. An overlay of the segmentation result is generated, which the user visually inspects for fit (Figure 1C). Further annotations are then drawn for each class or previous annotations are removed as necessary for any misclassifications, and the classifier is re-trained. This fine-tuning process is repeated until an experienced neuropathologist visually confirms the fit of the overlay of segmented classes with the original image. A final image is then generated that turns the classified pixels of each class into different colors, which allows for calculation of the area via counting the number of pixels of each color (Figure 1D, E). To quantify the percentage of amyloid deposition, the pixel count of amyloid is divided by the sum of pixel counts of amyloid, tissue, and calcifications. After segmentation of one image, the classifier and associated data can be saved and imported for use into a subsequent image.

**Figure 1.**
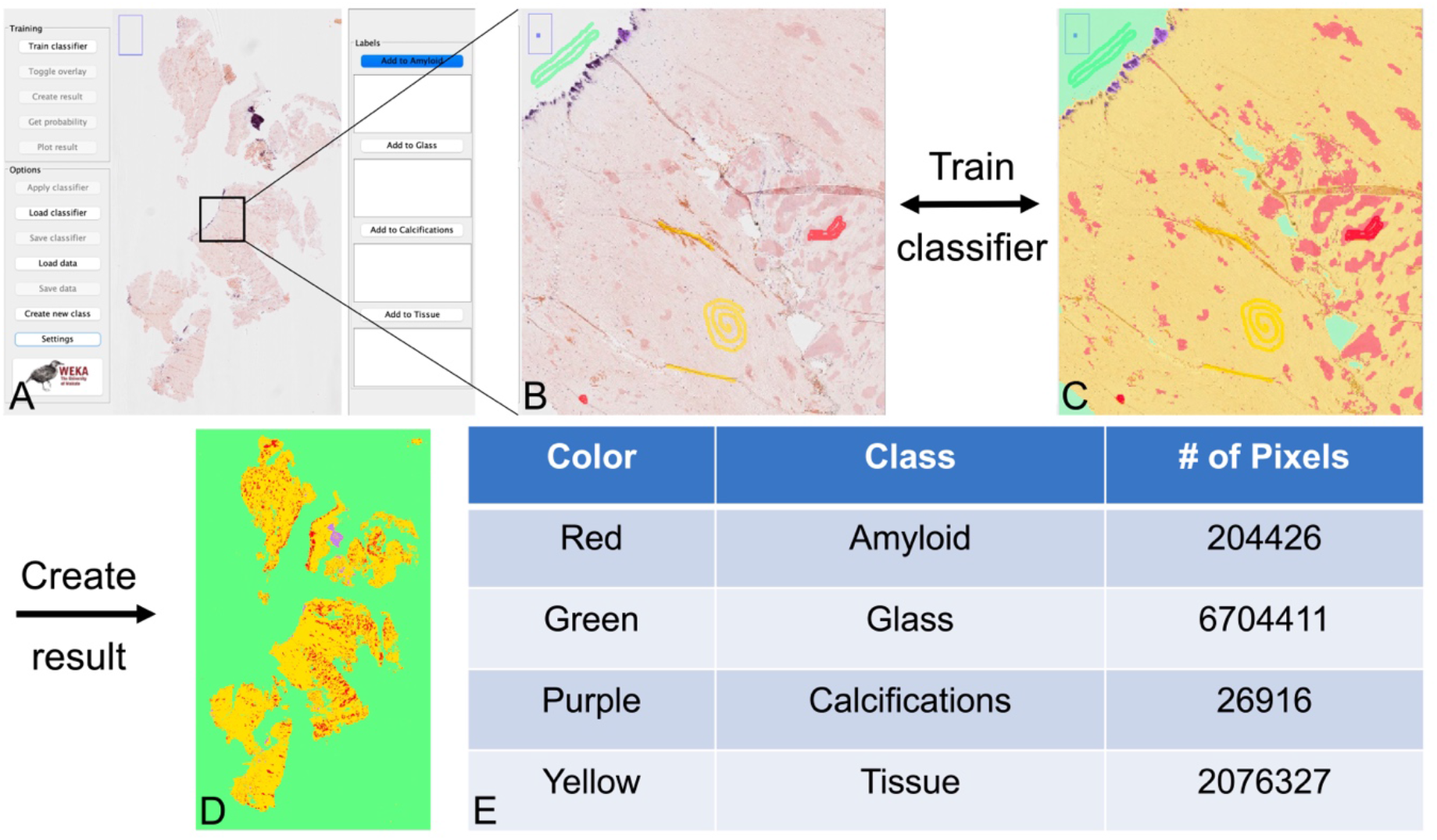
Segmentation and quantification of amyloid deposits using Trainable Weka Segmentation. A) An image of the ligamentum flavum specimen is imported into Fiji and opened within the TWS graphical user interface. B) A few manual annotations are drawn that correspond to each class of interest. C) The classifier is then trained to learn the characteristics of each class through these annotations and segments the rest of the image. An overlay is generated for the user to inspect the fit of the segmentation to the original image. Additional annotations may be added or prior annotations removed, re-training the algorithm as necessary until reaching desired fit. D) After satisfactory fit with the overlay, a final image is generated that assigns the pixels of each class to different colors. E) Calculation of the numbers of pixels in each class then allows for quantification of the total area of each component.

### Manual segmentation and color thresholding

Manual segmentation was performed using the freehand drawing tool in Fiji to individually trace around each amyloid deposit. An experienced neuropathologist visually confirmed each tracing to be accurate. Color thresholding was performed by using the color threshold function in Fiji to adjust the hue, saturation, and brightness levels of an image to capture the relevant salmon-pink color values that correspond to amyloid deposits [11]. The color values are adjusted individually for each histologic specimen until reaching the best achievable fit, also visually confirmed by an experienced neuropathologist.

### Statistical analysis

Statistical analysis was conducted using R version 4.1.1 and figures were produced using the package *ggpubr* [19,20]. Pearson correlation analysis was used to assess the association between machine learning segmentation and manual segmentation, as well as between color thresholding and manual segmentation. A p-value of < 0.05 was considered significant.

## RESULTS

### Use of TWS in single images with annotations and in automated quantification

Ten specimens of ligamentum flavum were obtained from spinal stenosis patients undergoing laminectomies and confirmed to have amyloid by Congo red staining demonstrating apple-green birefringence under polarized light, then analyzed to have a peptide profile consistent with ATTRwt via liquid chromatography tandem mass spectrometry (LC MS/MS, Mayo Clinic Laboratories, Rochester, MN, USA). The first ligamentum flavum image was imported into Fiji and opened within the TWS graphical user interface as described in the methods section and illustrated in Figure 1. Of note, only <5 human annotations per class were needed until the classifier reached a satisfactory level of fit through visual comparison to the original image. Figure 1C demonstrates the machine learning algorithm learning to accurately classify amyloid deposits and avoid misclassifying artifacts such as the large creases within the tissue that share the same salmon-pink color. The areas of amyloid in the final segmented image were confirmed to match well to the areas in the original image via close inspection by an experienced neuropathologist.

In addition to analysis of single histological images with annotations, TWS allows for automated quantification. The classifier trained by the first image can be saved and used in subsequent specimens of ligamentum flavum. However, the use of a TWS algorithm trained only with one image will likely be inaccurate, as Congo red staining across different specimens varies slightly in the shade of salmon-pink amyloid.

Therefore, the classifier trained with the first image was used in a subsequent set of training images and refined with additional annotations to fix any misclassifications. On additional images, the classifier was fine-tuned until an experienced neuropathologist visually confirmed the fit of the segmentations to the original images. This process was continued serially for the same classifier across a total of five training images. Figure 2A shows an enlarged side-by-side comparison of a LF specimen in its original form compared to its segmented image in one of the training specimens. Using this trained model from the initial five training scans, the classifier can then be used to automatically segment new images of LF without any further human annotations. The remaining five LF images were used as the application set, and TWS was used to quantify the amyloid deposits in these application images. Figure 2B shows an enlarged side-by-side comparison of a LF specimen in its original form compared to its segmented image in one of the application images segmented automatically by this process. Full-size images of the training and application sets with their corresponding segmented images are available in the appendix. The amyloid load, or percentage of amyloid deposition relative to the entire tissue, is calculated for all specimens and listed in Table 1.

**Figure 2.**
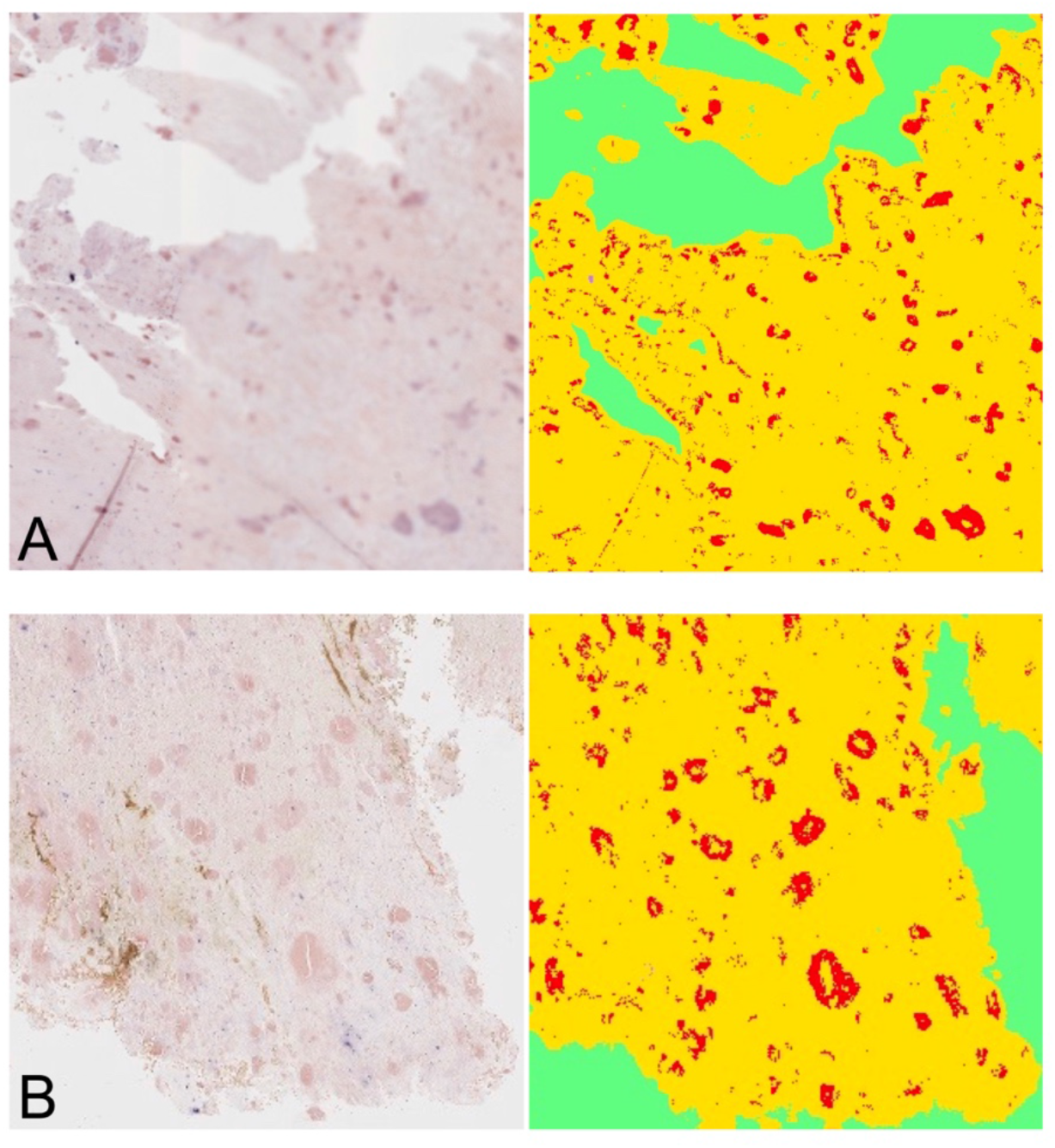
Comparisons of original and segmented ligamentum flavum images. A) Enlarged representative image from the training set. Left: original ligamentum flavum histology specimen stained with Congo red. Right: Segmented image generated by the TWS machine learning algorithm after training with annotations. B) Enlarged representative image from the application set. Left: original ligamentum flavum histology specimen stained with Congo red. Right: Segmented image generated by the trained TWS machine learning algorithm without additional annotations.

**Table 1.**
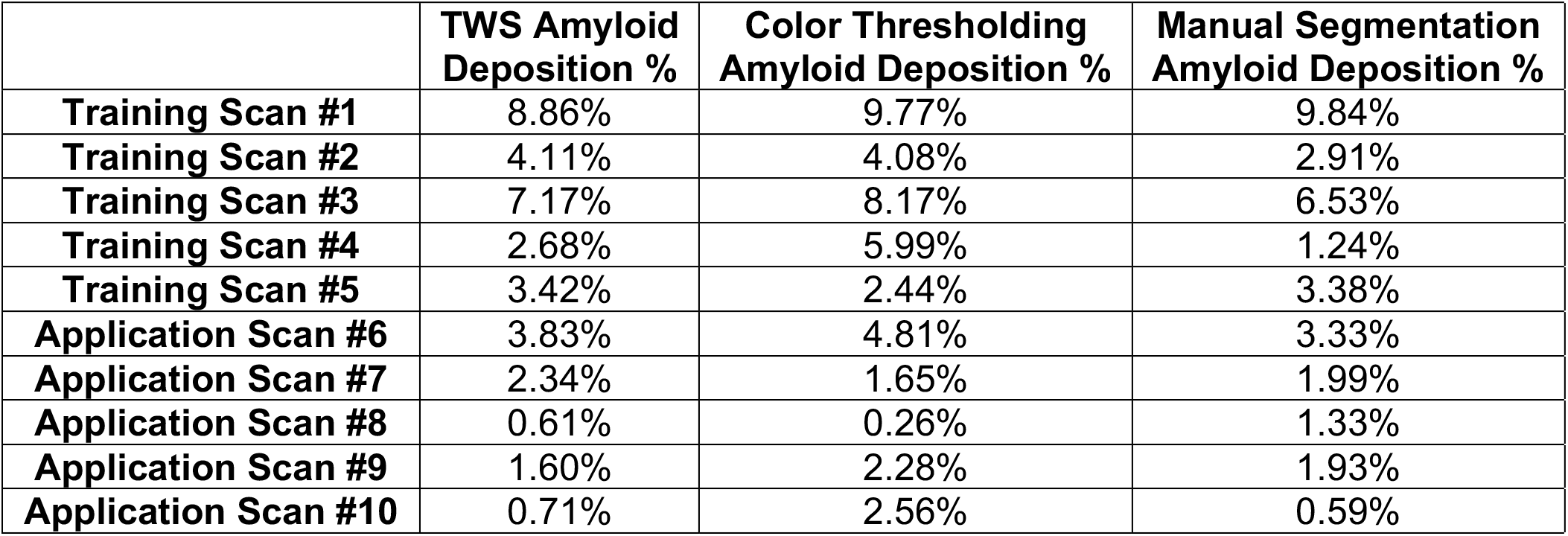
Quantification of amyloid load in ligamentum flavum scans from TWS, color thresholding, and manual segmentation. The percentage of amyloid deposition relative to the specimen in each scan was calculated by taking the number of pixels of amyloid divided by the sum of the pixels from amyloid, calcifications, and tissue. Amyloid load was calculated for each of the three methods used across all images.

### Comparison of TWS to color thresholding against manual segmentation

To compare TWS with other methods of quantifying amyloid in the ligamentum flavum, manual segmentation and color thresholding were also used to segment amyloid deposits in all ten images (Figure 3). Manual segmentation involved using the freehand drawing tool in Fiji to trace around each amyloid deposit under the supervision of an experienced neuropathologist (Figure 3C). Color thresholding involved using the function in Fiji to adjust pixel color values of hue, saturation, and brightness for each image to capture the relevant color values that correspond to amyloid deposits, also performed under supervision of an experienced neuropathologist (Figure 3D). The amyloid loads as quantified by these methods are calculated and listed in Table 1.

**Figure 3.**
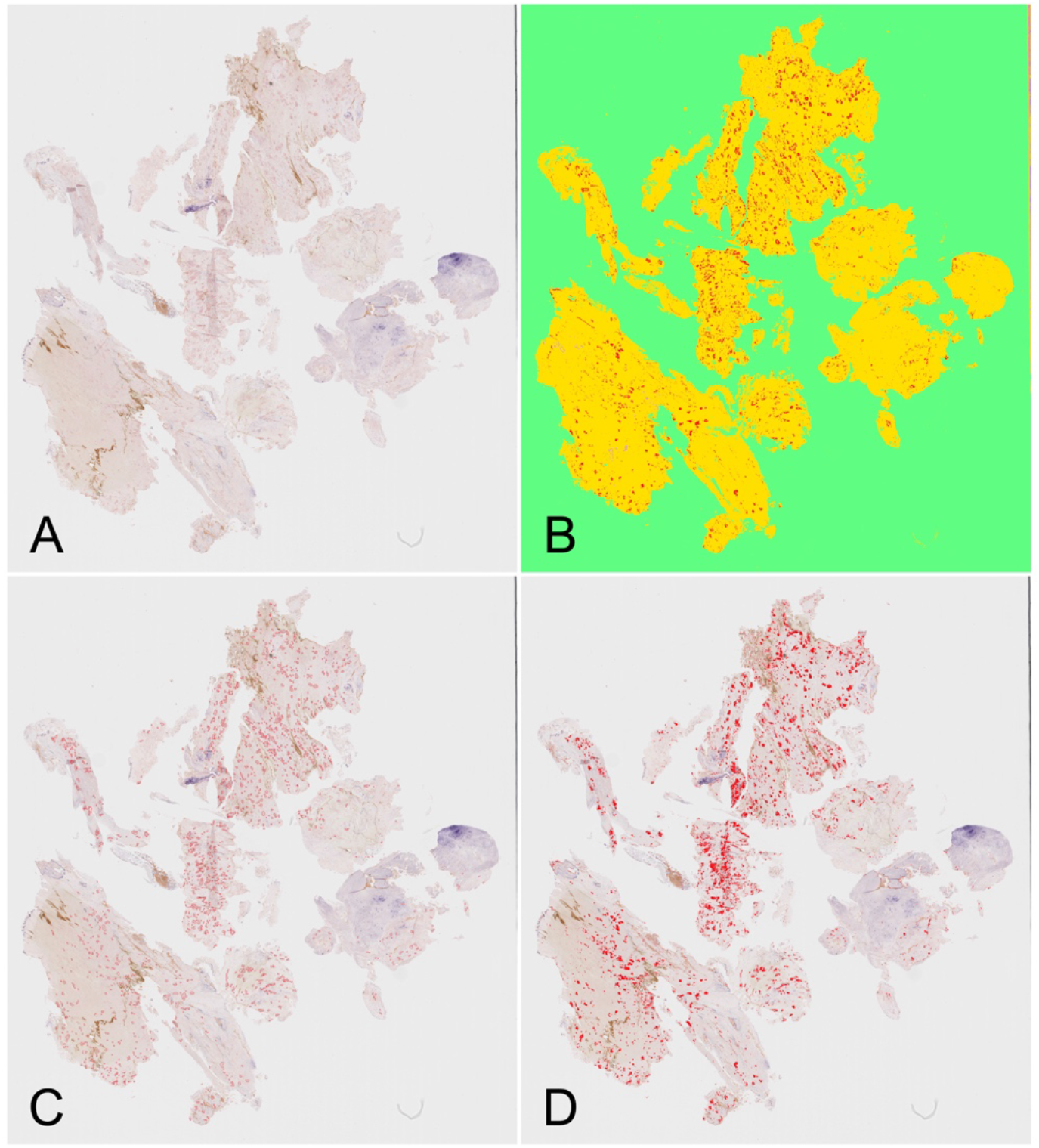
A single LF image from the application set segmented by the different methods of TWS, manual segmentation, and color thresholding. A) Full-size original image of ligamentum flavum. B) Segmented image after application of the trained TWS model. C) Segmented image via manual segmentation of amyloid deposits. D) Segmented image via color thresholding.

To assess the accuracy of TWS in segmenting and quantifying amyloid deposits, scatter plots compared how closely TWS correlated with manual segmentation (Figure 4). Manual segmentation is used as a gold standard comparator in both as it allows the neuropathologist the best control over identifying amyloid deposits [21]. The performance of TWS is compared to color thresholding through their correlations with manual segmentation. Two scatterplots were created, one of the set of five training scans (Figure 4A), and one of the other set of five application scans (Figure 4B). A least squares regression line was plotted, and Pearson correlation analysis performed to determine how closely either the TWS or color thresholding methods matched manual segmentation. In the training set, TWS is strongly correlated with the manual segmentation and achieved statistical significance (R = 0.98; p = 0.0033). Color thresholding is less strongly correlated with the manual segmentation and did not reach statistical significance (R = 0.78; p = 0.12). In the application set, TWS is also strongly correlated with the manual segmentation and achieved statistical significance (R = 0.94; p = 0.016). The color thresholding is also less correlated with the manual segmentation and did not reach statistical significance (R = 0.65; p = 0.23).

**Figure 4.**
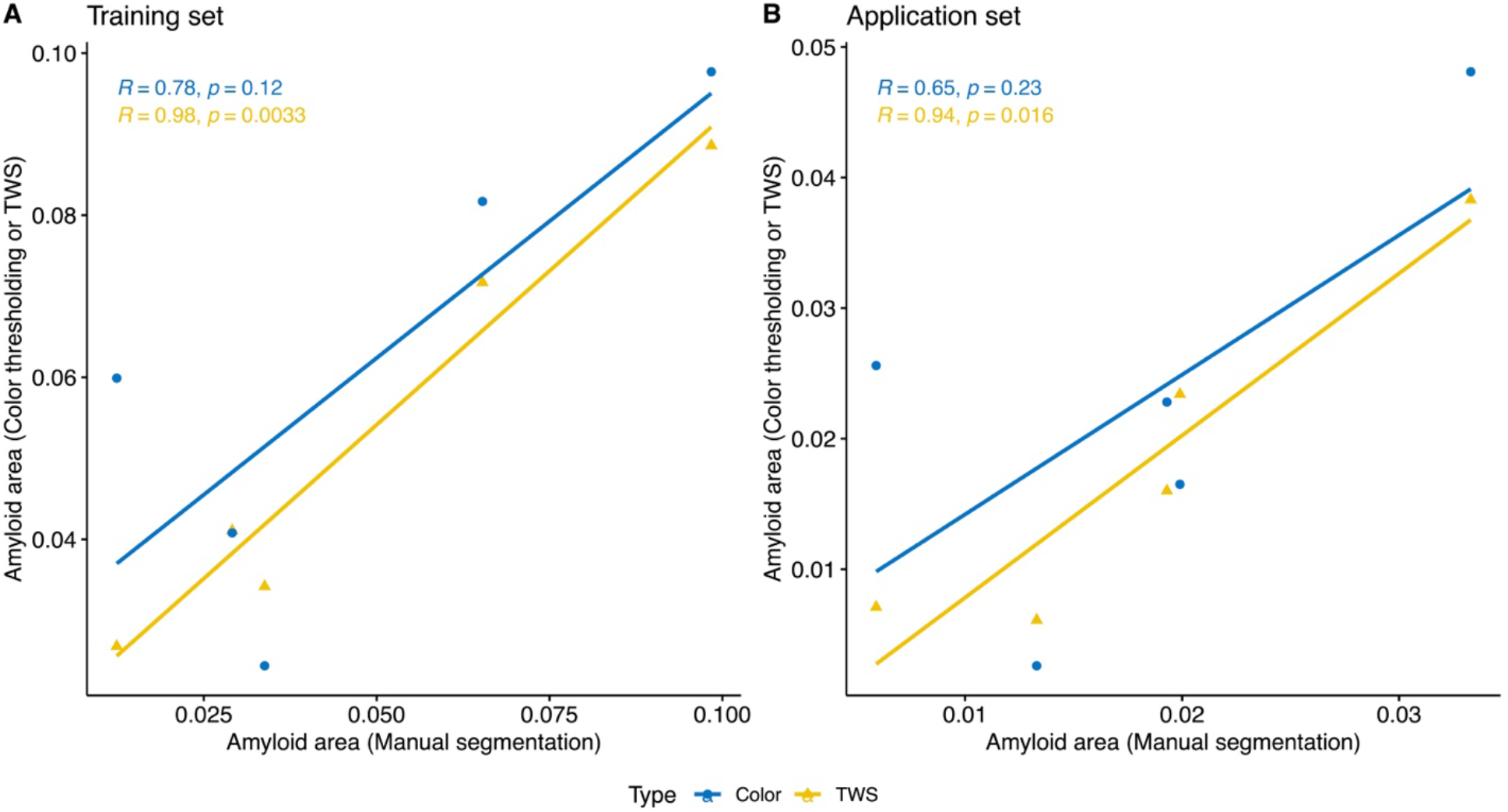
Comparison of TWS to color thresholding against manual segmentation. Scatter plots were made of the fractional area of amyloid deposition as quantified via TWS and color thresholding against that via manual segmentation. Pearson correlation analysis was used to compare the performance of TWS and color thresholding against manual segmentation. A) In the training set of images where TWS received user-directed annotations, the segmentation is strongly correlated with the manual segmentation (R = 0.98; p = 0.0033). The color thresholding is less strongly correlated with the manual segmentation (R = 0.78; p = 0.12). B) In the application set of images where TWS was automated without additional annotations, the machine learning segmentation is strongly correlated with the manual segmentation (R = 0.94; p = 0.016). The color thresholding is less correlated with the manual segmentation (R = 0.65; p = 0.23).

## DISCUSSION

Trainable Weka Segmentation is a powerful machine learning tool to quantify amyloid deposits in histological specimens of ligamentum flavum. Previous studies have been imprecise and/or not clearly described. Some utilized qualitative methods of visual inspection to distinguish amyloid deposits broadly into general categories rather than calculate a numerical area value [8–10]. Color thresholding has been well described, but we found the method to be subjective and difficult to reproduce [11]. Manual segmentation is theoretically the gold standard as it allows an experienced neuropathologist to maintain the best control over identifying amyloid deposits. However, in practical use this method is infeasible as it requires spending between 3-6 hours per specimen. This paper details a machine learning method to analyze amyloid deposits that overcomes the limitations of previous methods.

When used on single images with user-directed annotations, TWS proved to correlate closely with the gold standard of manual segmentation (Figure 4A). The classifier was sensitive at identifying amyloid and excluding artifact (Figure 1). In contrast, color thresholding correlated less well with manual segmentation and thus performed worse than TWS (Figure 4A). This is likely because color thresholding forces the user to overcount artifacts or undercount amyloid. Furthermore, it was remarkable how few annotations were required for TWS. In each image, usually <5 annotations per class were required to achieve a satisfactory fit by visual examination. In images with user-directed annotations, TWS quantifies amyloid with high accuracy and outperforms color thresholding.

Additionally, the TWS method allows for automation. Training the classifier across a training set of images generates a robust model that can be used to segment new application images without further human annotations. The training phase allows the classifier to capture the variations inherent in Congo red staining across different specimens, reliably segmenting amyloid despite the color variations. This can be seen in Figure 2B, where the application image as segmented by the trained classifier correctly identified amyloid deposits and excluded artifacts that shared the same salmon-pink color. In contrast, color thresholding frequently fails to distinguish artifact as it relies on the color values of each pixel, and many artifacts share the same color value as amyloid (Figure 3C). Figure 4B shows that in the set of application images, automatic segmentation via TWS still correlates well with results from manual segmentation, despite having no further human annotations. In the same set of application images, color thresholding continues to be less well correlated with manual segmentation (Figure 4B). These results highlight the promising potential of automatic segmentation via TWS.

This novel application of TWS to quantify amyloid in LF specimens has many advantages. Fiji and the TWS plugin are free to download and easily accessible by anyone with a regular laptop or computer, avoiding the expense of commercial machine learning algorithms. Also, the algorithm runs quickly, allowing users to quickly obtain segmentation results and rapidly re-train the algorithm during the fine-tuning process. This speed also enables use for high-throughput automation at scale for research and clinical use. Further broadening its accessibility is its easy-to-use graphical user interface that does not require special programming knowledge and its open-source distribution. These factors enable easy validation of data by other groups and encourages greater collaboration. If one master classifier were to be trained with many LF specimens and shared across different centers, this master classifier would allow researchers and clinicians to segment future samples without the need to train their own classifier.

TWS may open the door to future applications for research and clinical use. Our group hopes to use this method to explore the relationship of amyloid deposition in the LF with LF thickness and the progression of systemic amyloidosis. Precise quantification of amyloid load may enable correlation with clinical biomarkers and facilitate investigation into the histopathological development of amyloidosis. In this paper, we have only described the use of TWS to calculate the area of amyloid deposits, but this powerful technology can also study other characteristics of amyloid deposition such as the number of deposits, the differing sizes of deposits, and the presence of other notable substances such as calcifications. These other characteristics may have some currently unknown significance. Future research may also indicate a clinical use in patient care. One possible use might be as a screening tool to identify which spinal stenosis patients are at higher risk for developing cardiac amyloidosis and may benefit from further clinical follow-up. Outside of studying amyloid in the ligamentum flavum, the detailed workflow we described for using TWS may be easily applied to studying other pathologic processes of interest on any histological image.

This study has several limitations. One limitation is that machine learning is a “black box” and its segmentation process is not easily understood. We attempted to overcome this limitation by validating TWS in an application set of scans and comparing those results to other methods. With a limited training dataset, users will still likely need to visually check the segmentation of each image when automating, as the trained algorithm has not yet encountered all the possible variations among Congo red stains. Perhaps with a larger training dataset, there would be more confidence to reach the point at which visual confirmation would not be necessary. Another technical limitation is our use of only a few features for segmentation, which includes gaussian blur, difference of gaussians, and Sobel operator. The TWS plugin includes a wide range of features to use including edge detectors, texture filters, noise reduction filters, and membrane detectors [12]. Our decision to use just an important few was due to running the program on a standard laptop or desktop with a smaller amount of processing power, which may be addressed if powered through more advanced desktops or research computing clusters.

In summary, TWS is a powerful tool for accurately quantifying amyloid deposits in ligamentum flavum specimens. TWS with annotations can accurately identify amyloid and exclude artifacts. TWS can also be automated with a training subset of images and deployed in an application set without the need for further human adjustment. Training with five images was enough to create a robust model that was accurately deployed on five new images. TWS performed similarly well to the gold standard of manual segmentation and was superior to color thresholding. These results highlight this machine learning algorithm to be a quantitative, precise, unbiased, accessible, and high-throughput method to quantify amyloid in the ligamentum flavum.

## CONCLUSIONS

We present TWS as a novel, precise, objective, accessible, and high-throughput method to quantify amyloid deposits in histologic slides of ligamentum flavum stained with Congo Red. We find its efficacy to be comparable to that of manual segmentation and to outperform color thresholding. This robust method of quantifying amyloid in the ligamentum flavum may pave the way towards a variety of future research and clinical applications.

## Data Availability

Data produced in the present study are unavailable due to patient confidentiality.

---

**Appendix.**
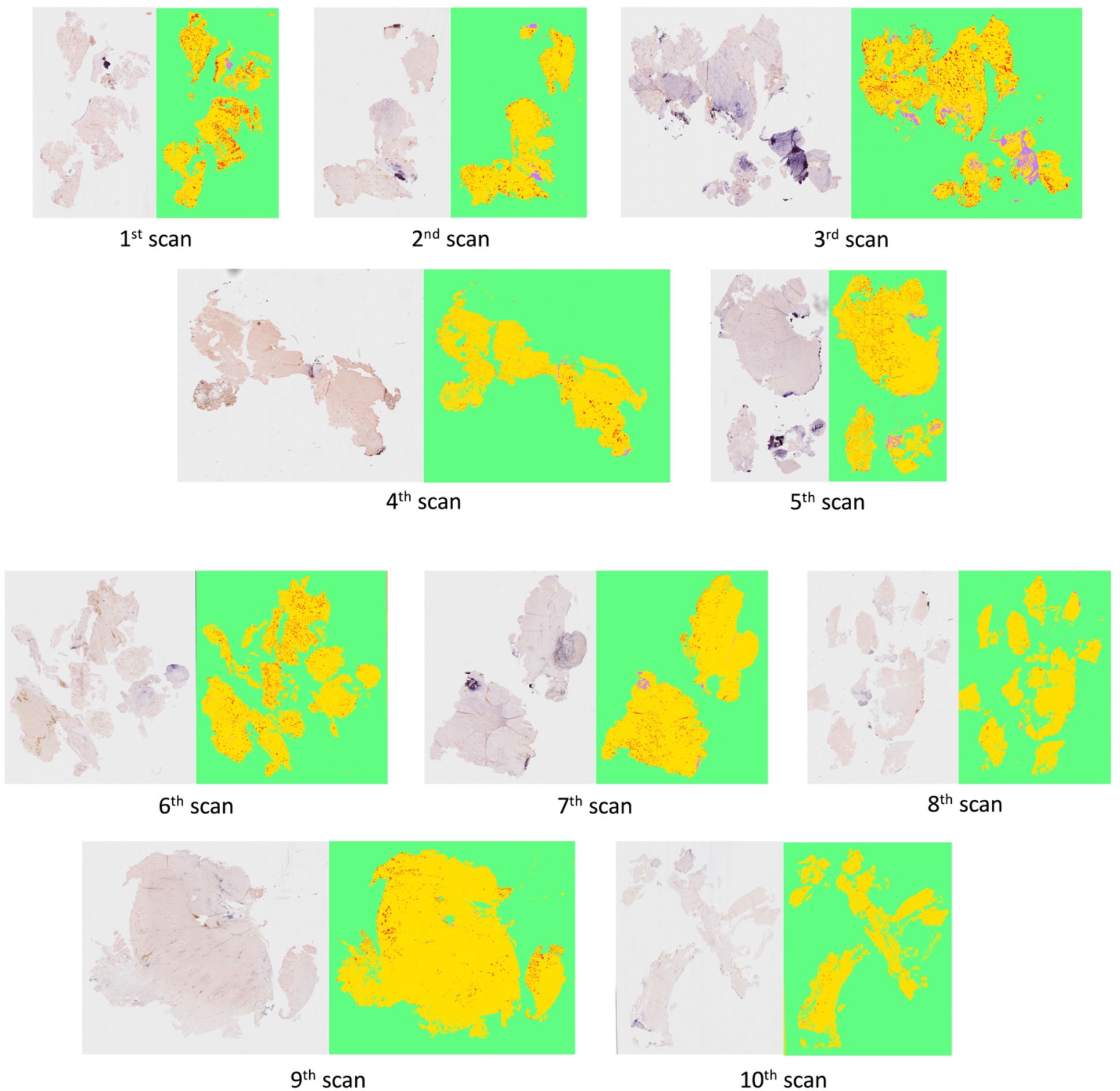
Full-size images of the original ligamentum flavum images compared with segmented images via TWS. Scans 1 to 5 are images from the training set. Scans 6 to 10 are images from the application set.

## Notes

**Funding:** The project was supported by the National Center for Advancing Translational Sciences, National Institutes of Health, Award Number TL1TR002546. The content is solely the responsibility of the authors.

### Competing Interest Statement

The authors have declared no competing interest.

### Funding Statement

Andy Y. Wang was supported by the National Center for Advancing Translational Sciences, National Institutes of Health, Award Number TL1TR002546. The content is solely the responsibility of the authors and does not necessarily represent the official views of the NIH.

### Author Declarations

The IRB of Tufts Medical Center gave ethical approval for this work (#13293).

